# Immune dysregulation caused by novel gain-of-function *UNC93B1* variant with enhanced antigen presentation

**DOI:** 10.64898/2026.03.03.26347379

**Authors:** Xu Han, Qintao Wang, Seza Ozen, Wei Dong, Yi Zeng, Ouyuan Xu, Seher Şener, Yunfei An, Li Guo, Ying Gu, Tingyan He, Jun Yang, Huanming Yang, Qing Zhou, Xiaomin Yu

## Abstract

UNC93B1 is a crucial chaperone protein for the trafficking of TLRs and regulates antigen presentation in dendritic cells (DCs), which activates downstream immune responses. Here, we identified a novel homozygous gain-of-function (GOF) *UNC93B1* variant in an early-onset lupus patient. The patient presented with elevated level of inflammation and auto-antibody, and organ damage. The *Unc93b1*^R95L/R95L^ transgenic mice also exhibited with autoimmune and autoinflammatory phenotypes. The transcriptional analysis revealed increased inflammation and elevated activation of DCs in the patient’s PBMCs and bone marrow-derived DCs (BMDCs) from *Unc93b1*^R95L/R95L^ mice. In addition to the selected TLR7/8 activation in previously reported *UNC93B1* GOF variants, the single-cell transcriptome and flow cytometry of splenocytes from *Unc93b1*^R95L/R95L^ mice demonstrated increased phagocytosis activity and T helper cell differentiation with altered ICAM and MHC signaling in DCs and T cells, respectively. These results suggest *UNC93B1* GOF variant enhances antigen presentation from DCs to T cells in the pathogenesis of immune dysregulation. Our study expands the pathogenic variants spectrum of *UNC93B1* and offers insight into the underlying mechanism of antigen presentation in immune dysregulation caused by UNC93B1 beyond its trafficking function of TLRs.

## Introduction

Immune dysregulation disorder is one characterization of inborn errors of immunity with heterogeneous manifestations including uncontrolled autoimmune, immunodeficiency, and autoinflammations caused by dysfunction of single gene in immune pathways^1,2^. Systemic lupus erythematosus (SLE) is a chronic systemic autoimmune disease with uncontrolled immune response that leads to multiorgan damage of varied severity^3^. Monogenic form of SLE predisposed to early-onset condition leads to worse clinical outcomes^4^ that could benefit from early genetic diagnosis. Till now, there are over 30 genes causing monogenic lupus been reported^5^.

Toll-like receptors (TLRs) are one of the earliest decisive components of innate immune activation^6^. Endosomal TLRs, including TLR3, TLR7, TLR8, TLR9, are transmembrane receptor recognizing pathogen-associated molecular patterns like various nucleic acids (NAs) and initiate downstream inflammatory pathways like NF-κB and type I interferon (IFN)^7^: TLR3 recognizes double-stranded RNA^8^, TLR7/8 recognizes single-stranded and short double-stranded RNA^6^, and TLR9 recognizes CpG single-stranded DNA^9^. Proper transportation of endosomal TLRs from endoplasmic reticulum (ER) to endosome is crucial for discrimination of foreign NAs and host NAs^6^. The expression of endosomal TLRs is specific in particular cells: TLR3 mainly expressed in conventional dendritic cells (cDCs), TLR7/9 mainly expressed in plasmacytoid dendritic cells (pDC) and B cells, while TLR8 mainly expressed in cDCs and monocytes^10-12^. TLRs also regulate the process of antigen presentation in dendritic cells (DCs) with tubulation and maturation of phagocytic compartment for major histocompatibility complex (MHC) processing^13,14^. The dysregulated signaling of endosomal TLRs have been recognized as a cause of immune dysregulation presenting with inflammation, immunodeficiency and lupus^15,16^

Unc-93 homolog B1 (UNC93B1), an ER-resident multi-transmembrane protein, could bind TLRs in the ER and traffic the endosomal TLRs from ER to their compartments and plays an important role in the immune response induced by endosomal TLRs activation^17,18^. Activated TLRs signaling triggers inflammatory response and could enhance the priming of antigen-presenting cells, such as DCs, to T cells^19^. Recently, a series of *UNC93B1* gain-of-function (GOF) variants were reported as pathogenic, predisposing to early-onset SLE and arthritis with systemic inflammation and autoimmune manifestation in patients and mice models through UNC93B1-TLR7/8 axis excluded TLR3 and TLR9^20^. However, the communication between immune cells during the development of lupus caused by GOF in *UNC93B1* is still not clear.

DCs, a type of antigen-presenting cells, are sentinels of immune system that activated by sensing pathogen or damage associated molecular pattern and regulating T cell homeostasis through signaling like major histocompatibility complex class (MHC) and ICAM^21,22^ with engulfment and phagocytosis of pathogens^23,24^. MHC-I signaling present endogenous antigens to activate cytotoxic T lymphocytes (CTLs) and MHC-II signaling present extracellular antigens to activate CD4^+^ T cells^25,26^, while ICAM signaling strengthens the immune synapse of T cells engaged by DCs^27,28^. Dysregulated UNC93B1 lead to affected antigen presentation through MHC signaling in murine DCs^18^. The orchestration of DCs plays pivotal role in the homeostasis of immune tolerance and pathogenic autoimmune response^29^.

Here, we demonstrate a novel GOF *UNC93B1* variant R95L in an early-onset SLE patient and give an insight into the alteration of DCs function in its immune response and antigen presentation to T cells during the development of immune dysregulation caused by GOF UNC93B1.

## Material and Methods

### Ethic approval

This study was conducted under the review and approval by the Institutional Review Boards at the Children’s Hospital of Zhejiang University School of Medicine in China with approval number: 2021-IRB-172. Written Informed consent was obtained from the patient and the patient’s parents.

### Mice

Generation of the R95L mutation mice model (C57BL/6J) of *Unc93b1* gene (ENSMUST00000162708.7) is via CRISP/Cas9 technology. The oligo donor DNA is: 5’CAGCCTTGCCGTGAGCTTGTTTAGTTGTTCTGAGCCAGACTGATTAGAGCTCTCTACGATGCTCCCTGTCCCCAGGCCTCCTGCAGATGCAACTGATCCTGCACTATGATGAGACCTACCTCGAGGTGAAGTATGGCAACATGGGGCTGCCGGACATCGATAGCAAGATGCTGATGGGTATCAACGTGACGCCTATCGCTGCCCTGCTCTACACACCTGTGCTCATCAGGTG CCAAACTTC-3’. The sgRNA is 5’-GCCATACTTCACCTCTCTGT-AGG-3’.

Animal experimentation was approved by the Institutional Animal Care and Use Committee of Zhejiang University with approval number: 2024-208. Both male and female mice were used and their genders are indicated in the figures or figure legends. Within genotypes, mice were randomly allocated in all experiments. Data collection and analysis were not performed blind to the conditions of the experiments, except for histopathology. No animals or data points were excluded except contaminated samples. Littermate are used as controls for comparison with *Unc93b1*^R95L/R95L^ mice. Serum levels of *Unc93b1*^R95L/R95L^ mice were assessed every two weeks from postnatal day 21 until the phenotype stabilized.

### Whole exome sequencing

Genomic DNA from the patient and family members were isolated from peripheral blood using Maxwell RSC Whole Blood DNA Kit (Promega, AS1520). Whole exome sequencing (WES) was performed on genomic DNA on Illumina NovaSeq 6000 platform with 150 bp paired-end reads and 100× average coverage. FASTQ files are mapped to human hg38 genome with bwa (version 0.7.17) mem algorithm. Variant calling is performed by using GATK standard workflow. ANNOVAR (2019Oct24) was used to annotate variants. Variants that appeared in gnomAD, Kaviar, dbSNP, and an in-house database were filtered out. Variants were further filtered by homozygous inheritance.

### Antibodies

For flow cytometry experiments, V500 Rat Anti-Mouse CD45 (30-F11) (561487), BV421 Hamster Anti-Mouse CD3e (145-2C11) (562600), FITC Rat Anti-Mouse CD4 (RM4-5) (553046), BV650 Rat Anti-Mouse IL-17A (TC11-18H10) (564170), BV711 Hamster Anti-Mouse CD279 (PD-1) (744547), and PE-CF594 Rat Anti-Mouse CD185 (CXCR5)(2G8) (562856) antibodies were purchased from BD Biosciences. eFluor™ 450 Anti-Mouse T-bet (48-5825-82) was purchased from Invitrogen.

### RNA extraction and quantitative PCR

TRIzol reagent (Invitrogen, 15596026) and RNeasy Mini kit (Qiagen, 74104) were applied to extract total RNA from cultured cells and murine tissues. One microgram of RNA was reverse-transcribed using the PrimeScript RT Reagent Kit with gDNA Eraser (Perfect Real Time) (Takara, RR047A). Gene expression analyses were performed using 2X Universal SYBR Green Fast qPCR Mix (ABclonal, RK21203) on ROCHE 480II. Relative mRNA expression was analyzed by the ΔΔ*C*_t_ method and normalized to *ACTB* (for human samples) or *Actb* (for mouse samples).

### RNA sequencing

RNA sequencing libraries were generated using the NEBNext Ultra RNA Library Prep Kit for Illumina (New England Biolabs) following the manufacturer’s protocol. The libraries were then sequenced on an Illumina NovaSeq platform to obtain high-throughput sequencing data. STAR (version 2.7.5) and featureCounts (version 2.0.1) were utilized to reads-mapping and counting. For differential expression analysis, the DESeq2 (version 1.44.0) and clusterProfiler (version 4.12.0) R package was employed. Pathway scores in differential expression analysis of RNA sequencing data are summing of z-scores of each gene in the gene set.

### Single-cell RNA sequencing

For human PBMCs, samples were captured and barcoded with a 10x Genomics Chromium console, while the library of murine spleen samples were constructed by MGI DNBelab C-TaiM4, following the instruction of manufacturers. Sequencing was performed on Illumina Novaseq and MGI DNBSEQ-T7 for human PBMC and murine samples, respectively. Sequencing data was processed with cellranger (10X Genomics, version 6.1.1) and DNBC4tools (MGI, version 2.1.1) for Novaseq and T7 data, respectively. Downstream quality control, processing and differential expression analysis were performed by Seurat package (version 4.2.0) and harmony package (version 1.2.0) in R.

### ELISA and CBA

Levels of cytokines IL-6, IL-10 in human serum were determined by Cytometric Bead Array (BD Bioscience, 558276, 558274). All data were analyzed by FCAPArray V3 software (BD Biosciences). For detecting autoantibodies in murine plasma, coating antigens (1 μg DNA or RNA) were diluted in Coating Buffer and added to an ELISA plate, then incubated overnight at 4 °C. The plate was blocked with Diluent Buffer for 1 hour at room temperature. Samples and standards were diluted in Diluent Buffer and incubated for 2 hours at room temperature. Detection antibodies were added and incubated for 1 hour, followed by HRP conjugate (Beyotime, A0216) for 30 minutes, both at room temperature. TMB substrate (Beyotime, P0209) was added and incubated for 30 minutes, then stop solution (Beyotime, P0215) was added. Absorbance was measured at 450 nm using a microplate reader.

### Immunohistochemistry

Freshly dissected murine spleens, kidneys, livers and lungs were fixed in 4% paraformaldehyde (Sigma-Aldrich, P6148) for 16 hours at 4 °C, embedded with paraffin and stained with hematoxylin/eosin (H&E) or periodic acid-Schiff (PAS). The tissue slides were scanned using Aperio ScanScope XT scanner (Leica).

### Flow cytometry analysis

Single-cell suspensions of murine splenocytes were generated by plunger end of syringe with filtering of 100 and 40 µm strainers in PBS with 2% FBS. The filtrates were centrifuged at 350 × g for 5 minutes. The cell pellet was suspended in 5 mL RBC lysis buffer, incubated for 4 minutes at 25 °C and then mixed with 10 mL of PBS with 2% FBS. The splenocytes were resuspend in PBS after centrifugation at 350 × g for 5 minutes. Whole blood cells were collected from heart blood and incubated with 10 mL RBC lysis buffer for 5 minutes at 25 °C, and then mixed with 20 mL PBS with 2% FBS. The whole blood cells were resuspended in PBS after centrifugation at 350 × g for 5 minutes.

For flow cytometry analysis, 1 × 10^6^ cells were incubated with Fixable Viability Stain 440UV (BD Biosciences, 566332) for 15 minutes at room temperature protected from light. Washed twice with fluorescence-activated cell sorting (FACS) buffer (0.5% BSA in PBS) and stained with cell-surface markers for 1 hours on ice. For fixation and permeabilization, 1.6% PFA and 40% methanol were used. The cells were then washed twice and stained with intracellular antibodies for 1 hours on ice. All events were acquired on Cytek Aurora and analyzed by SpectroFlo and FlowJo (version 10.8.1).

### Statistical analysis

Statistical analyses were performed using GraphPad Prism (version 10.1.2). All values were as mean ± SEM. Statistical significance were assessed with the unpaired two tailed Student’s *t* test for two groups and one-way ANOVA analysis for 3 groups. Differences were considered as statistically significant if *P* < 0.05. Permutation test based on hypergeometric distribution and Bonferroni correction was used in the *P* value calculation of GSEA significance and performed by clusterProfiler (version 4.12.0).

## Results

### Identification of novel homozygous *UNC93B1* GOF variant

The proband presented with fever, fatigue, discoid, maculopapular rashes, and photosensitivity with an early age of onset (**Fig. 1A**). Immunophenotyping of the patient’s immune cells reveals increased number of total T cells (90 %, reference: 56-84%) and expanded CD4^+^ T cell population (62 %, reference: 31-52%) in blood. Immunologic study of the patient demonstrates positive autoantibodies like anti-nuclear antibody, anti-double-stranded DNA antibodies, anti-Smith antibody, and anti-ribonucleoprotein. Laboratory tests of the patient show reduced level of C3 (0.33 g/L, reference: 0.8-1.6 g/L) and C4 (0.05 g/L, reference: 0.15-0.4 g/L) and elevated erythrocyte sedimentation rate. Taken together, the patient was diagnosed as early-onset SLE evaluated by SLEDAI and SLEDAI-2k scoring system. The patient presently received daily administration of 5 mg prednisolone, 150 mg hydroxychloroquine, and mycophenolate mofetil.

**Figure 1.**
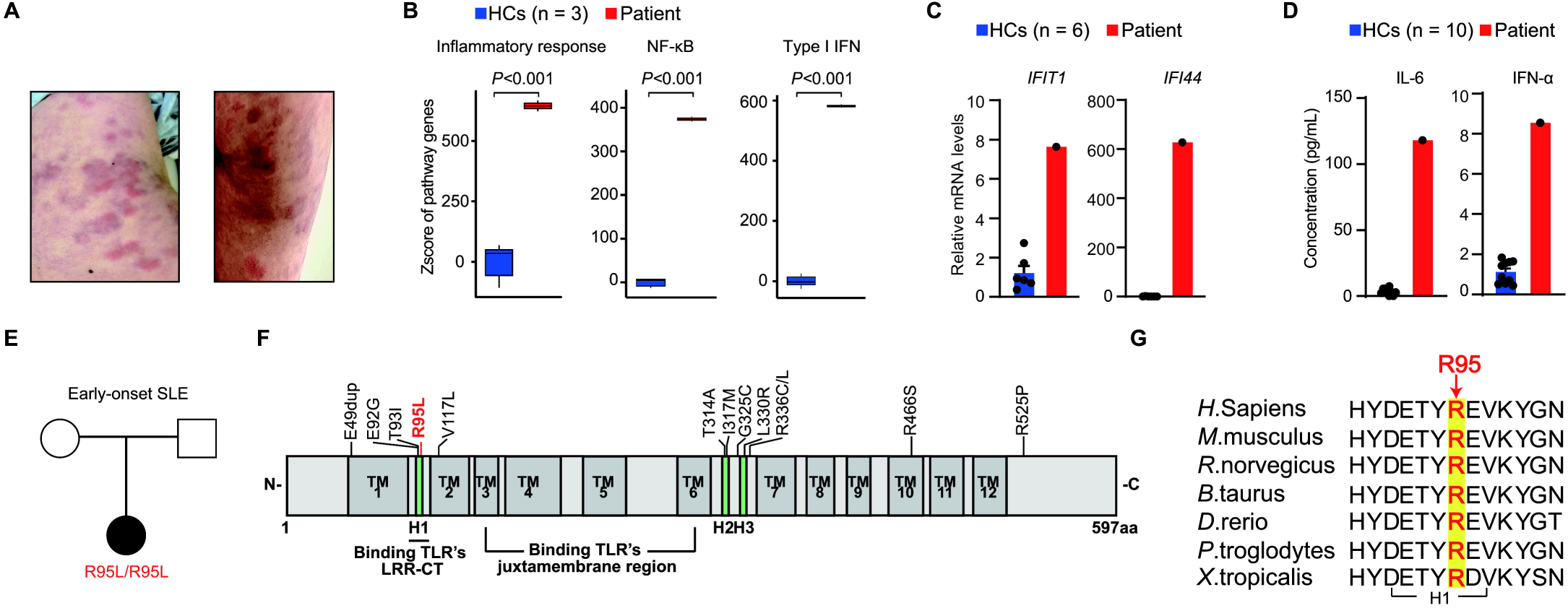
Activated inflammatory signaling in an early-onset SLE patient with *UNC93B1* R95L variant. (A) Clinical images displaying skin rashes on the patient’s back (left) and hip (right). (B) Comparison of Z-scores of indicated gene sets like inflammatory response, NF-κB, and type I IFN between the patient and healthy controls (n = 6). HCs, healthy controls. Unpaired Student’s *t* test is used in statistics. Boxes show the median (center line), 25th and 75th percentiles (upper and lower hinges), and whiskers extend to 1.5× the interquartile range. (C) Transcriptional level of interferon-related genes in the patient analyzed by qPCR compared to healthy controls (n = 6). HCs, healthy controls. (D) Serum level of inflammatory cytokines in the patient detected by cytometric bead array compared to healthy controls (n = 10). HCs, healthy controls. (E) Pedigree of the patient with a homozygous variant NM_030930:c.284G>T, p.R95L in *UNC93B1*. (F) Structure diagram of UNC93B1 protein with known GOF pathogenic variants and R95L variant marked. TM, transmembrane domain. (G) The evolutionary conservation of the arginine at position 95 in UNC93B1 across 7 species.

To evaluate the differential inflammatory response in the patient, we performed bulk RNA sequencing on the peripheral blood mononuclear cells (PBMCs) from the patient and healthy controls. The analysis on differential expressed genes suggested pathway scores are elevated on inflammatory response, NF-κB, and type I IFN signaling in the patient compared to healthy controls(**Fig. 1B**). The expression profile of NF-κB and type I IFN pathway was also upregulated compared to healthy controls (**Fig. S1A**). Gene set enrichment analysis (GSEA) showed top-ranked upregulated genes enriched on type I IFN and TNF signaling in the patient (**Fig. S1B**). Quantitative PCR (qPCR) of the PBMCs from the patient validated the increased transcription of crucial genes on type I IFN pathway, like *IFIT1* and *IFI44* (**Fig. 1C**), while cytometric bead array (CBA) assay on the serum samples from the patient revealed pronounced increased production of proinflammatory cytokines like IL-6 and IFN-α in the patient(**Fig. 1D**). These results demonstrated dysregulated immune response in the patient and indicated dominant contribution of NF-κB and type I IFN pathways in the patient’s immune dysregulation manifestation.

Whole-exome sequencing identified a novel homozygous variant in *UNC93B1* (c.284G>T, p. R95L) (**Fig. 1E**). The R95 residue is located on the H1 domain of UNC93B1 interacting with TLRs^30^ as well as known pathogenic variants like E92G and T93I (**Fig. 1F, table S1**) and is highly conserved across species(**Fig. 1G**). The variant had a CADD score of 35, supporting a predicted deleterious effect on protein function. The R95L in *UNC93B1* is potentially pathogenic to the patient’s immune dysregulation manifestation.

### Activation of TLR7/8 signaling caused by R95L variant leading to immune dysregulation

To test the effect of the *UNC93B1* R95L variant in the pathogenesis of autoimmune pathology and dysregulated immune response, we generated mice model carrying heterozygous or homozygous *Unc93b1* R95L mutation using CRISPR/Cas9 technology. Compared to wild type (WT) mice, the R95L variant has no impact on the expression of Unc93b1 in both *Unc93b1*^+/R95L^ and *Unc93b1*^R95L/R95L^ mice (Fig. S2A-C). Compared to wild type mice, *Unc93b1*^R95L/R95L^ mice exhibited significant splenomegaly (**Fig. 2A**). The serum level of autoantibodies like anti-double-stranded DNA or RNA (anti-dsDNA or anti-dsRNA) antibodies was significantly increased in female *Unc93b1*^R95L/R95L^ mice (**Fig. 2B**), while both male and female *Unc93b1*^+/R95L^ mice showed slightly increased autoantibody signal.

**Figure 2.**
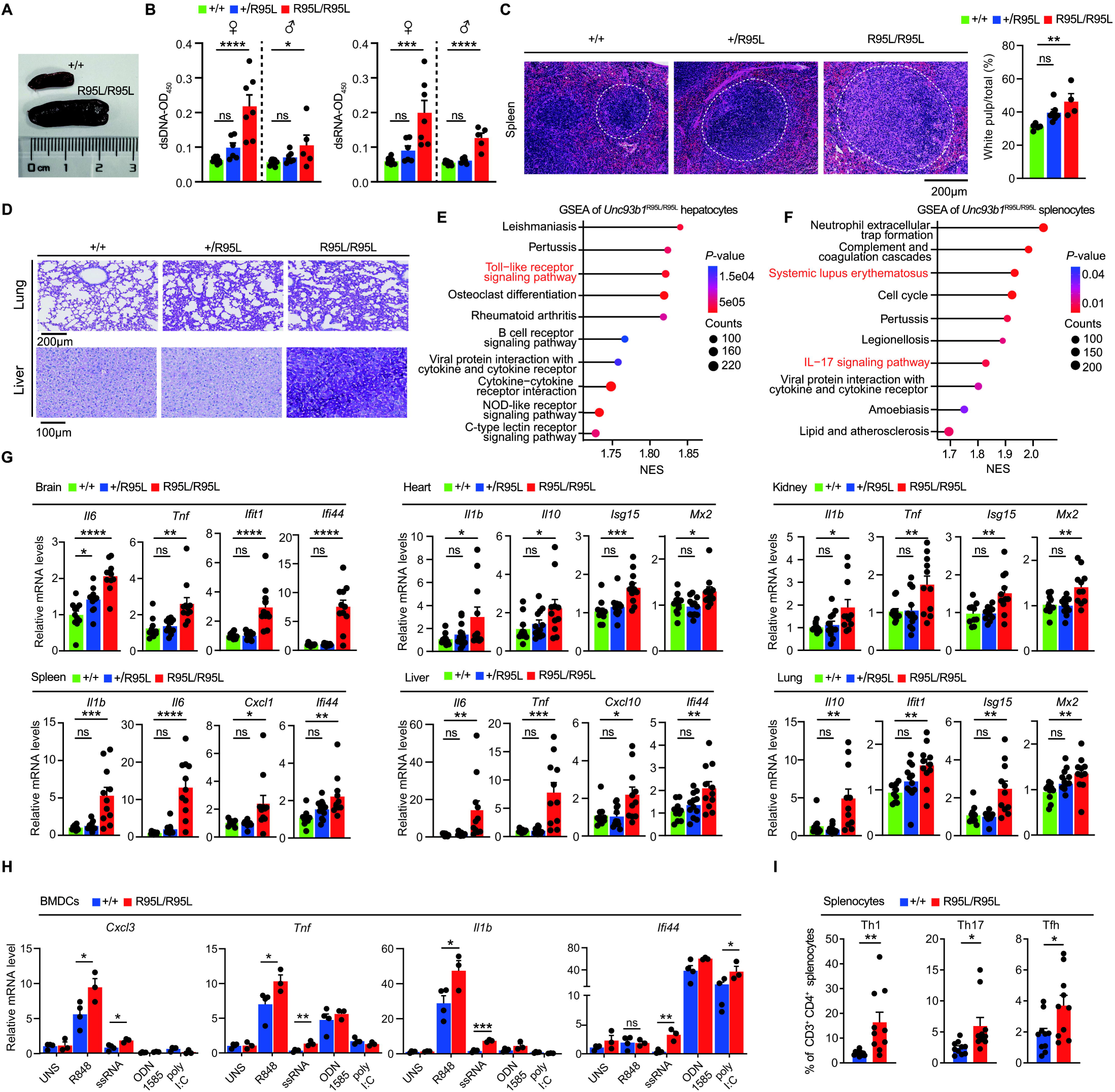
R95L mutation drives immune dysregulation pathology in *Unc93b1*^R95L/R95L^ mice. (A) Splenomegaly of *Unc93b1*^R95L/R95L^ mice compared to WT mice. (B) Autoantibody levels of *Unc93b1*^+/+^, *Unc93b1*^+/R95L^, *Unc93b1*^R95L/R95L^ mice detected by ELISA (*Unc93b1*^+/+^, female: n = 10, male: n = 9; *Unc93b1*^+/R95L^, female: n=6, male: n = 7; *Unc93b1*^R95L/R95L^, female: n = 7, male: n = 5). SEM and one-way ANOVA with Tukey’s post hoc analysis is used in statistics; *, *P* < 0.05; **, *P* < 0.01; ns, non-significance. (C) Hematoxylin and eosin staining of spleen tissue from *Unc93b1*^+/+^, *Unc93b1*^+/R95L^, and *Unc93b1*^R95L/R95L^ mice (left). White pulp boundary is circled by white dot line. Normalized area of white pulp is calculated (*Unc93b1*^+/+^, n = 5; *Unc93b1*^+/R95L^, n = 7; *Unc93b1*^R95L/R95L^, n = 4). SEM and one-way ANOVA with Tukey’s post hoc is used in statistics; *, *P* < 0.05; **, *P* < 0.01; ***, *P* < 0.001; ****, *P* < 0.0001; ns, non-significance. (D) Periodic acid-Schiff staining of lung (upper) and liver (bottom) tissue from *Unc93b1*^+/+^, *Unc93b1*^+/R95L^, and *Unc93b1*^R95L/R95L^ mice. Reduced alveolar spaces and thickened septa indicate lung damage, while excessive glycogen storage with darker purple staining indicates liver damage. (E) Enrichment of upregulated pathways of differentially expressed genes in hepatocytes from the *Unc93b1^R95L/R95L^* mice based on GSEA, compared to *Unc93b1*^+/+^ mice. NES, normalized enrichment score. *P* value is calculated by one-sided hypergeometric distribution test and adjusted by Bonferroni correction. Permutation times of GSEA is set to 1000 as default. (F) Enrichment of upregulated pathways of differentially expressed genes in splenocytes from the *Unc93b1^R95L/R95L^* mice based on GSEA, compared to *Unc93b1*^+/+^ mice. NES, normalized enrichment score. *P* value is calculated by one-sided hypergeometric distribution test and adjusted by Bonferroni correction. Permutation times of GSEA is set to 1000 as default. (G) qPCR analysis of inflammatory genes among different organs of *Unc93b1*^+/R95L^ and *Unc93b1*^R95L/R95L^ mice (n = 10 mice in groups except heart and liver; n = 11 mice in heart and liver of *Unc93b1*^+/+^ group, kidney of *Unc93b1*^+/R95L^ group and all *Unc93b1*^R95L/R95L^ group; n = 12 mice in all *Unc93b1*^+/R95L^ group except kidney). SEM and one-way ANOVA with Tukey’s post hoc is used in statistics; *, *P* < 0.05; **, *P* < 0.01; ***, *P* < 0.001; ****, *P* < 0.0001; ns, non-significance. (H) qPCR analysis of genes related to NF-κB and type I IFN pathway in BMDCs from *Unc93b1*^R95L/R95L^ mice (n = 4 mice in *Unc93b1*^+/+^ group; n = 3 mice in *Unc93b1*^R95L/R95L^ group) treated with corresponding TLRs ligands (R848, 1μM; ssRNA, 2μM; ODN1585, 2μM; poly(I:C), 10 μg/mL). SEM and unpaired two-tailed Student’s *t* test is used in statistics. *, *P* < 0.05; **, *P* < 0.01; ***, *P* < 0.001; ns, non-significance. (I) Population of T helper cell in splenocytes of *Unc93b1*^R95L/R95L^ mice analyzed by flow cytometry, compared to *Unc93b1*^+/+^ mice (n = 10 in each group). SEM and unpaired two-tailed Student’s *t* test is used in statistics. *, *P* < 0.05; **, *P* < 0.01. Th, T helper; Tfh, T follicular helper.

To investigate the effect of R95L variant on the organ of *Unc93b1*^R95L/R95L^ mice, we tested the difference of pathology and immune response between *Unc93b1*^R95L/R95L^ mice and their littermate controls. Hematoxylin and eosin staining in spleen tissue revealed enlarged white pulp indicating splenic lymphoid hyperplasia in *Unc93b1*^R95L/R95L^ mice, which suggests altered immune response in spleen (**Fig. 2C**). With periodic acid-Schiff staining, we also observed reduced alveolar spaces and thickened septa in lung tissue of *Unc93b1*^+/R95L^ or *Unc93b1*^R95L/R95L^ mice (**Fig. 2D, upper)**, and excessive glycogen storage in liver tissue of *Unc93b1*^R95L/R95L^ mice (**Fig. 2D, lower**), which indicate organ damage in lung and liver, respectively.

To find the exact signaling in the pathology of *Unc93b1*^R95L/R95L^ mice, we also performed bulk RNA sequencing on the hepatocytes and splenocytes from *Unc93b1*^R95L/R95L^ mice and WT mice. The analysis of the differentially expressed genes indicated an elevated pathway score of NF-κB pathways in both liver and spleen of *Unc93b1*^R95L/R95L^ mice (**Fig. S2D**). Compared to WT mice, GSEA on transcriptome also revealed top-ranked upregulated genes were enriched in immune signaling, like TNF, NF-κB, TLR pathway in hepatocytes, and IL17, SLE signaling, TNF, and TLR pathway in splenocytes, respectively, in *Unc93b1*^R95L/R95L^ mice (**Fig. 2E and F, Fig. S2E**). To evaluate the level of systemic inflammation caused by R95L variant, we tested the expression level of essential genes on NF-κB and type I IFN pathway in different organs from *Unc93b1*^R95L/R95L^ mice. Compared to WT mice, the qPCR analysis of various organs, including brain, heart, kidney, spleen, liver, and lung, reveals a significantly upregulated systemic inflammation signal in *Unc93b1*^R95L/R95L^ mice(**Fig. 2G**). In summary, *Unc93b1*^R95L/R95L^ mice present with immune dysregulation, while *Unc93b1*^R95L/R95L^ mice were asymptomatic, but their pathology differed from WT mice.

Hyper-activated endosomal TLR signaling could lead to immune dysregulation phenotype with excessive activation of downstream inflammatory signaling like NF-κB and type I IFN signaling and autoimmune manifestation like SLE^15,31^. According to the essential regulatory effect of UNC93B1 on endosomal TLRs, we investigated the effect of *UNC93B1* R95L variant on the response of downstream signaling of endosomal TLRs. Previously reported GOF *UNC93B1* variants exhibited selective activation of TLR7/8 but not TLR3/9 signaling^20^. By stimulated with specific TLR agonists, we tested alteration of immune response in bone marrow-derived dendritic cells (BMDCs) from *Unc93b1*^R95L/R95L^ mice under stimulation of corresponding TLR ligands. Compared to WT mice, the qPCR analysis demonstrated significantly upregulated expression of genes on NF-κB and type I IFN signaling responding to TLR7 ligands than TLR3 or TLR9 ligands (**Fig. 2H**). The UNC93B1 and TLR7 signaling play a pivotal role in the regulation of antigen presentation in DCs^18,19,32^, which suggests an essential role of UNC93B1 and TLRs in phagocytosis and antigen processing. Besides, Activated DCs migrate to lymphoid nodes and prime T cell differentiation through antigen presentation^33,34^. Consistent with previous research^35^,the expansion of T helper cells is confirmed by cytometry of splenocytes from *Unc93b1*^R95L/R95L^ mice (**Fig. 2I**), suggesting the activation of antigen presentation from DCs to T cells in the spleen.

These results demonstrated the selective activation of *UNC93B1* R95L variant in TLR7 signaling and confirmed its pathogenesis to immune dysregulation with manifestation of autoimmune, systemic inflammation, and organ damage. The selectively activated TLR7 signaling in BMDCs and the expansion of T helper cells in splenocytes suggest an activated antigen presentation and T cell priming of DCs in *Unc93b1*^R95L/R95L^ mice.

### R95L variant enhanced antigen presentation of DCs in murine splenocytes

To investigate the suggested activation of DCs and T cells and explore the specific cell population contributing to the dysregulated immune response, we performed single-cell transcriptional analysis to the splenocytes from *Unc93b1*^R95L/R95L^ mice and WT mice. There are 26 cell populations identified based on unsupervised clustering(**Fig. 3A, Fig. S3A**). GSEA on splenocytes from *Unc93b1*^R95L/R95L^ mice demonstrated significantly upregulated enrichment on phagocytosis-related signaling, such as lysosome and efferocytosis, in DCs from *Unc93b1*^R95L/R95L^ mice (**Fig. 3B**), which suggested enhanced engulfment and phagocytosis in the process of antigen presentation. We also observed enhanced T helper differentiation signaling and activated SLE signaling in splenic T and B cells from *Unc93b1*^R95L/R95L^ mice, respectively (**Fig. 3C**). With the AddModuleScore method, the expression level of gene sets also confirmed the enhanced signaling of T helper differentiation and antigen presentation in T cells (**Fig. 3D, Fig. S3B**) and DCs (**Fig. 3E, Fig. S3C**) from *Unc93b1*^R95L/R95L^ mice, respectively. Besides, in splenocytes from *Unc93b1*^R95L/R95L^ mice, we observed that the score on NF-κB pathway elevated, especially DCs and T cells population, while the B cell population showed elevated signal in immunoglobulin production pathway (**Fig. S3D-E**). These results suggested the promoted T cell differentiation and activated DCs in *Unc93b1*^R95L/R95L^ mice, which explained the pathology from *Unc93b1*^R95L/R95L^ mice with upregulation of inflammatory and autoimmune signaling.

**Figure 3.**
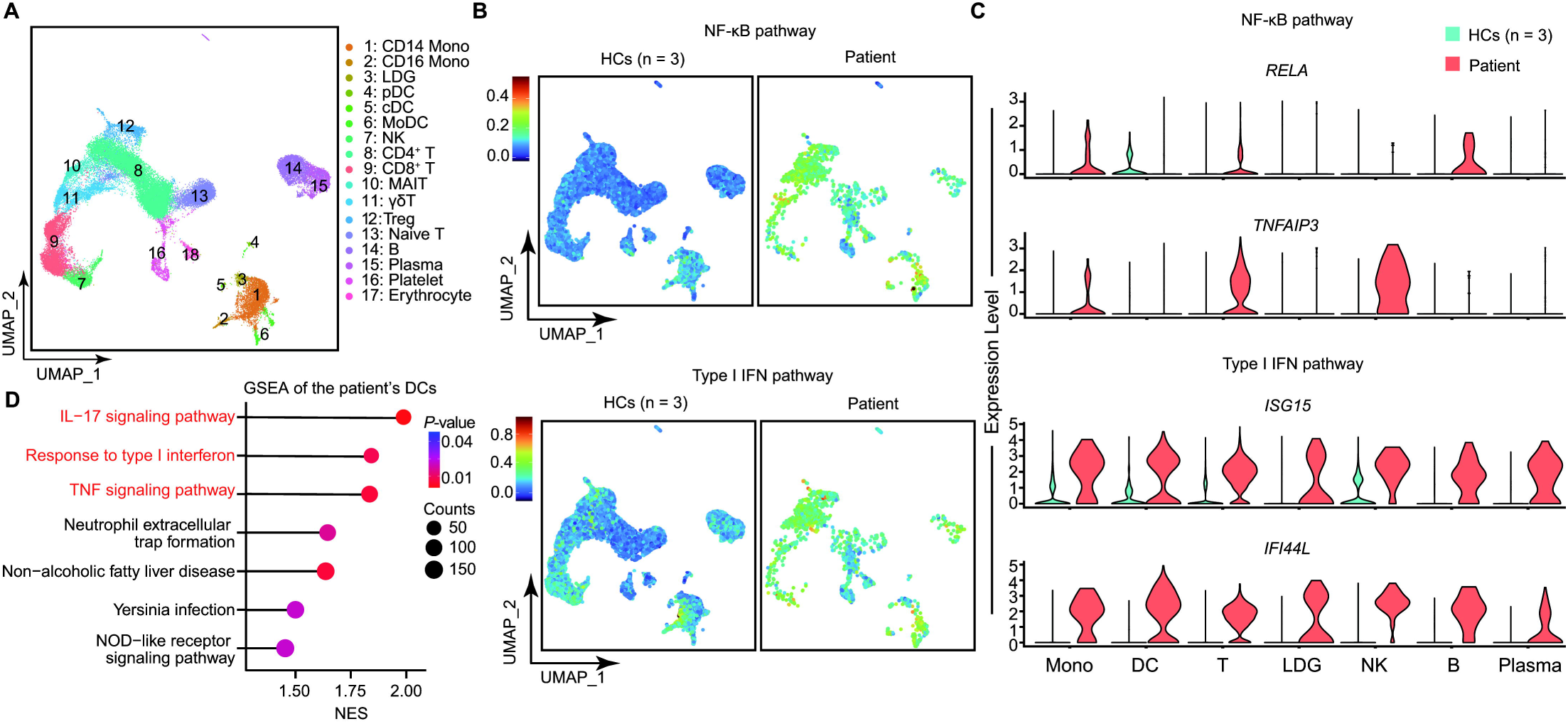
Single-cell analysis of splenocytes from *Unc93b1*^R95L/R95L^ mice suggests enhanced antigen presentation caused by *Unc93b1* R95L mutation. (A) UMAP visualization and marker-based annotation of 28 cell subtypes from splenocytes of *Unc93b1^R95L/R95L^* and *Unc93b1^+/+^* mice (n = 3 mice per group). NKT, natural killer T cell; γδT, gamma-delta T cell; Treg, regulatory T cell; MZ B, marginal zone B cell; Fo B, follicular B cell; DC, dendritic cell; MDP, myeloid dendritic progenitor. (B) GSEA analysis of phagocytosis-related pathway in the splenic DCs from *Unc93b1*^R95L/R95L^ mice compared to *Unc93b1*^+/+^ mice (n = 3 in each group). NES, normalized enrichment score. *P* value is calculated by one-sided hypergeometric distribution test and adjusted by Bonferroni correction. (C) GSEA analysis of autoimmune and T helper differentiation pathway in the splenic B and T cells from *Unc93b1*^R95L/R95L^ mice compared to *Unc93b1*^+/+^ mice (n = 3 in each group). NES, normalized enrichment score. *P* value is calculated by one-sided hypergeometric distribution test and adjusted by Bonferroni correction. (D) Expression level of genes in T helper cells differentiation in *Unc93b1^R95L/R95L^*mice. The expression level is calculated by the Seurat Addmodulescore method and mapped to the UMAP plot. Detailed statistical significance between WT and *Unc93b1^R95L/R95L^* mice is listed in the Fig. S4B. (E) Expression level of genes in antigen presentation in *Unc93b1^R95L/R95L^* mice. The expression level is calculated by the Seurat Addmodulescore method and mapped to the UMAP plot. Detailed statistical significance between WT and *Unc93b1^R95L/R95L^* mice is listed in the Fig. S4C. (F) Differential taking or sending signaling of splenic DCs from *Unc93b1*^R95L/R95L^ or *Unc93b1*^+/+^ mice. Positive and negative value indicates difference of signaling strength in *Unc93b1*^R95L/R95L^ compared to *Unc93b1*^+/+^ mice. The color shows specific or shared signaling in the indicated set. (G) ICAM signaling from splenic DCs to T cells in *Unc93b1*^R95L/R95L^ and *Unc93b1*^+/+^ mice. Treg, T regulatory; CTL, cytotoxic T lymphocyte. (H) Expression levels of ligands and receptors in ICAM signaling. (I) MHC signaling from splenic DCs to T cells in *Unc93b1*^R95L/R95L^ and *Unc93b1*^+/+^ mice. Treg, T regulatory cells; CTL, cytotoxic T lymphocyte.

Conventional DCs (cDCs) and plasmacytoid DCs (pDCs) are main DC subsets in spleen, who exert a role on T cell priming, including the differentiation of T helper cells and effector T cells, and type I IFN production, respectively^36^. The activated pDCs could also present NA antigens and activate CD4^+^ T cells^37^. Given that NKT and γδ T cells participated in the antigen presentation through a CD1-mediated mechanism^38,39^, but not MHC, we focused on the communication between DCs and CD4^+^/CD8^+^ T cells. The analysis on differential signaling communication showed specific enhanced outgoing ICAM signaling and altered MHC-II signaling from splenic DCs of *Unc93b1*^R95L/R95L^ mice (**Fig. 3F**). The expression of essential genes on ICAM signaling (**Fig. 3G**) and detailed signaling direction and strength of ICAM from splenic DCs to T cell (**Fig. 3H**) revealed the contribution of cDC2 population in the enhancement of ICAM signaling. The detailed communication of MHC signaling form splenic DCs to T cells demonstrated an apparently augmented MHC-I presentation to CTLs and enhanced MHC-II signaling to CD4^+^ T cells from cDC2 and pDCs (**Fig. 3I**).

These results indicate promoted antigen presentation to CTLs and enhanced priming of CD4^+^ T cells in splenocytes from *Unc93b1*^R95L/R95L^ mice, which described a picture of exact DCs subsets like cDC2 and pDCs in the increased antigen presentation in the immune dysregulation caused by *UNC93B1* R95L variant.

### Single-cell analysis confirmed the activated inflammation of DCs and T cells in the patient’s PBMCs

To confirm the activation of DCs and T cells in the patient and explore exact cell populations contributing to the alteration of immune response, we analyzed single-cell transcriptional profile of the patient’s PBMCs. There are 17 cell populations identified based on unsupervised clustering (**Fig. 4A, Fig. S4A**). Based on the AddModuleScore method of Seurat^40^, scores on NF-κB pathway were elevated in overall cell populations, especially myeloid cells and T cells, while type I IFN pathway was activated in most populations (**Fig. 4B-C, Fig. S4B-C**). Compared to healthy controls, GSEA demonstrated that DCs and T cells from the patient showed upregulated genes enriched on inflammatory signaling, including IL17, type I IFN, and TNF signaling (**Fig. 4D, Fig. S4D**).

**Figure 4.**
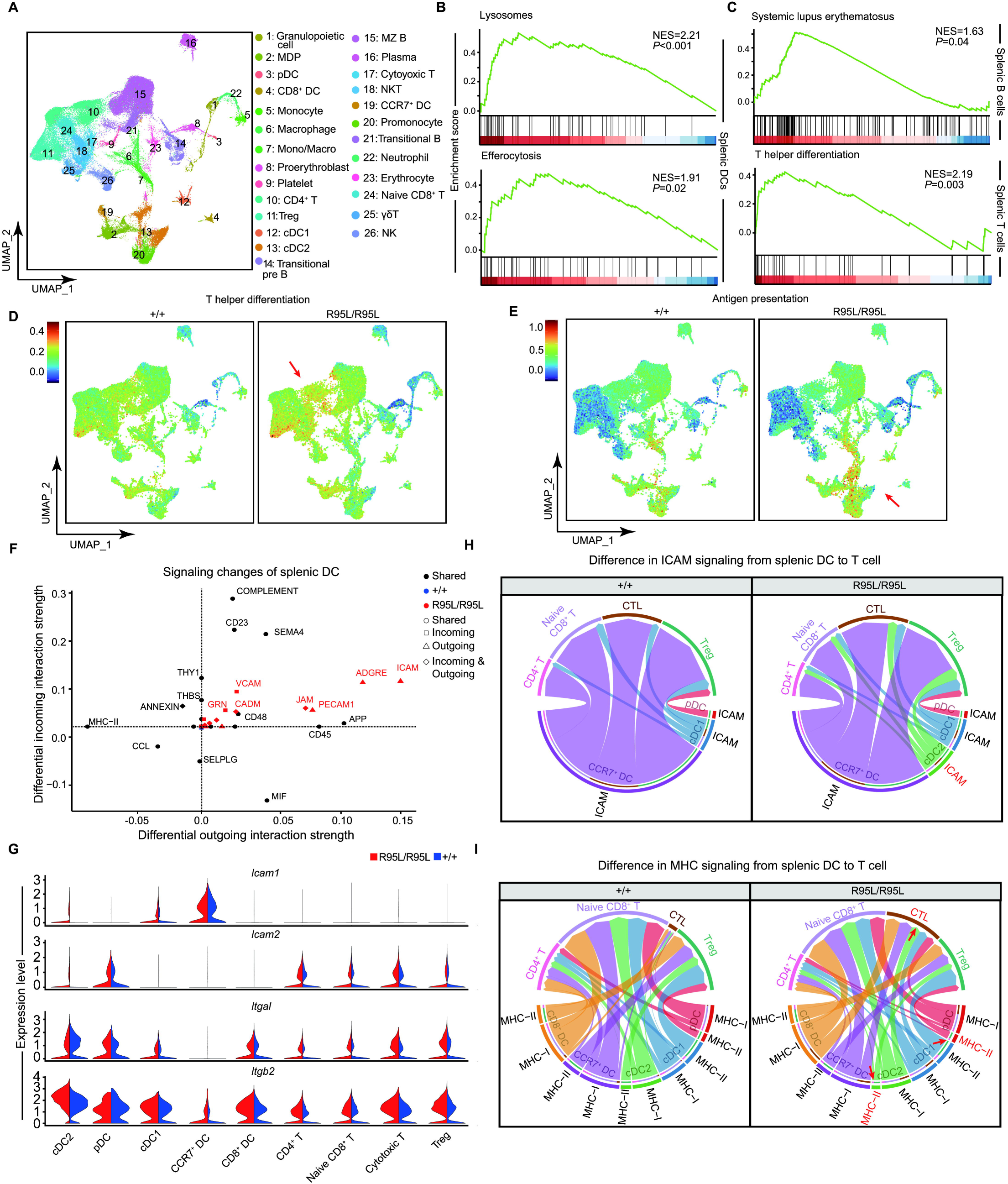
Transcriptomic analysis of the patient’s PBMC reveals inflammatory signal alteration across immune cells. (A) Uniform manifold approximation and projection (UMAP) visualization and marker-based annotation of 17 cell subtypes from the patient and healthy controls (HCs, n = 3). LDG, low-density granulocytes; pDC, plasmacytoid dendritic cells; cDC, conventional dendritic cells; MoDC, monocyte-derived DC; MAIT, mucosal-associated invariant T cell; NKT, natural killer T cell; γδT, gamma-delta T cell; Treg, regulatory T cell; MK, megakaryocytes. (B) Upregulated expression of genes in NF-κB (upper) and type I IFN (bottom) pathway gene set of MSigDB in the patient. The expression level is calculated by the Seurat Addmodulescore method. Detailed statistical significance between healthy controls and *the* patient is listed in the Fig. S3B-C. (C) Expression levels of essential genes in NF-κB and type I IFN pathways in various immune cells among the patient and healthy controls (HCs, n = 3). (D) Enrichment of upregulated pathways of differentially expressed genes in the patient’s DCs based on GSEA. NES, normalized enrichment score. *P* value is calculated by one-sided hypergeometric distribution test and adjusted by Bonferroni correction. Permutation times of GSEA is set to 1000 as default.

These results suggested that *UNC93B1* R95L variant arouse the elevated downstream inflammatory pathways including IL17 signaling, NF-κB signaling, and type I IFN signaling, which suggested the activation of DCs and T cells in the manifestation of the patient with *UNC93B1* GOF variant.

## Discussion

In conclusion, our results identify a novel homozygous GOF *UNC93B1* variant, expand the spectrum of immune dysregulation, and emphasize the regulatory function of UNC93B1 on antigen presentation in the pathogenesis of immune dysregulation beyond TLRs trafficking. TLR7-rendering *UNC93B1* variant D34A leads to hyperresponsivity of TLR7, facilitates the differentiation of CD4^+^ T cells to T helper cells, which eventually leads to systemic inflammation, autoimmunity, and organ damage like splenomegaly, and hepatic necrosis^17^. Given the specific expression of endosomal TLRs in DCs (TLR3/8 in cDCs, while TLR7/9 in pDCs)^10^, the research on skewed endosomal TLRs signaling regulated by UNC93B1 in different DCs and the UNC93B1-regulated antigen presentation in phagocytes will be attractive. Aside from DCs, other myeloid cells, such as monocytes^41^, are also antigen-presenting, which may also hypothetically be an effector in the patient’s pathogenesis. Accompanied by the myeloid cells and lymphocytes, the homeostasis orchestrated by TLRs, UNC93B1 and MHCs is important in both innate and adaptive immunity and will inspire the research of the overlapped autoinflammatory and autoimmune manifestations in immune dysregulation patients in the future.

In previous research, both mono- and bi-allelic of *UNC93B1* GOF variant drives the pathogenesis of the patient’s phenotype. We noticed that patients with mono-allelic variants appear to be more prone to an asymptomatic state(**Fig. S5A**), while the phenotype caused by mono- or bi-allelic UNC93B1 variants has no obvious predisposition. Variants on UNC93B1 residues, such as V117, E92, and R336, implicated a gene dosage effect in previous studies^35,42,43^. Though the mechanism is still elusive, decreased protein stability in the UNC93B1 mutant^43^, weakening the suppressing interaction between UNC93B1 and TLR7 with syntenin-1^35,43-45^, hypothetically implies an accumulative role of damaging alleles in the differential penetrance of hetero/homozygotes of GOF *UNC93B1*. Here, though the *Unc93b1*^+/R95L^ mice in our analysis exhibited partial autoimmune pathology in organ, they did not present with autoimmune manifestation, compared to *Unc93b1*^R95L/R95L^ mice. Besides, our patient’s parents, who carry heterozygous *UNC93B1* R95L variant, also have no clinical complaints, consistent with healthy GOF *UNC93b1* carrier parents of the patient in previous research^43^. Hence, carriers of *UNC93B1* R95L variant are not symptomatic. Nevertheless, more attention still should be paid to carriers of GOF *UNC93B1* variants in clinical practice. Besides, we also observed that the onset-age of patients with bi-allelic *UNC93B1* variants seems more concentrated on early age, while the onset-age of patients with mono-allelic UNC93B1 variants could be delayed till the teenage (**Fig. S5B**), which suggests a worse severity in patients with bi-allelic *UNC93B1* GOF variants.

There are molecules participating in the regulation of UNC93B1-mediated endosomal TLRs transportation to their destined compartment, the endosome. Associated with adaptor protein (AP) 4, UNC93B1 delivers TLR7 from the ER to the endosome, while the trafficking of TLR9 uses AP-2 to transport TLR9 from the ER to the plasma membrane and destines to the endosome^45^. Different from TLR7, UNC93B1 dissociated with TLR9 on the endosome for its activation^46^. Similar to TLR9, the release from UNC93B1 is also crucial for downstream signaling activation of TLR3^46^. Therefore, these molecules assisting the UNC93B1-TLR axis are potentially pathogenic to monogenic SLE. Recently, defect on another endosomal molecule, PLD4, who maintain the homeostasis of endosomal NAs, also led to an accumulation of NAs within the endosome and triggered SLE^31^. Therefore, the homeostasis in the endosome could also be an interesting topic in the pathogenesis of monogenic SLE.

Given the well response of JAK inhibitor in the monogenic SLE caused by deficiency of endosomal molecule PLD4 in mice model^31^, it could also be a potential treatment for our patient according to the enhanced type I IFN signaling driven by activation of TLR7/8 caused by GOF UNC93B1. Besides, by regulating the acidification of the endosome, hydroxychloroquine could inhibit endosomal TLR activation^47^ and is used as an approved treatment of SLE^48^, which could be administrated on our patient as well. These precise treatments could overcome the limitation of lacking specificity in traditional SLE therapy^49^ combining with genetic diagnosis. Thereby, the clinical trial of other inhibitors to the crucial molecules on TLR7/8 signaling, including Myd88., IRAK1/2/4, and TRAF6, could be encouraged in the future. However, some patients may be allergic to specific drugs, and the treatment of baricitinib may fail due to biological resistance in some individuals^50^. Besides, adverse events can occur when a long-term treatment spanning years is administered, including infections for baricitinib, and ophthalmological or cardiac complications for HCQs^51-53^. Therefore, a tailored therapy with customized treatment for specific individuals is necessary. A treatment with alternative drugs, such as ruxolitinib, another JAK inhibitor, or anifrolumab, a monoclonal antibody for type I IFN that is suitable for SLE patients, could be considered for patients with refractory manifestations or patients who are allergic or resistant to a specific drug. In our patient, the elevation of serum IL-6 was also linked to the upregulated inflammatory state. IL-6 inhibitors, like tocilizumab and siltuximab, have been used for the treatment of escalated IL-6 signaling^54-56^.

Taken together, our findings offer a view of augmented antigen presentation signaling caused by GOF variant in *UNC93B1* in the pathogenesis of immune dysregulation disorders. Next step to evaluate the driving effect of GOF *UNC93B1* variant in different region of UNC93B1 will not only help to disentangle the detailed functions of UNC93B1 domains, but also allow the drafting of criteria for assessing the severity of clinical outcomes of different GOF *UNC93B1* variant combinations, which facilitate precise medicine to the patients.

## Data availability

All data are available in the main text or the Supplemental materials upon request.

## Acknowledgments

We thank the patient and the unaffected controls for their support during this research study. We thank the core facility of the Life Sciences Institute, Zhejiang University and core facilities of Zhejiang University Medical Center/Liangzhu laboratory for technical assistance.

## Author contributions

X.Y., Q.Z. designed the study, directed, and supervised the research. X.H., Q.W., and S.O. contributed equally. Q.W., Y.Z., and O.X. performed experiments. X.H., W.D., Y.G., and T.H. analyzed the data. Q.Z., S.O., S.S., A.Y., L.G., J.Y., and H.Y. enrolled the patient, collected, and interpreted clinical information. X.Y., X.H., and Q.W. wrote the manuscript, with input from others. All authors contributed to the review and approval of the manuscript.

