## Supplementary material for "Immune dysregulation caused by novel gain-of-function *UNC93B1* variant with enhanced antigen presentation": UNC93B1_Supplementary_Material v1.1.pdf

**Supplementary Figure**

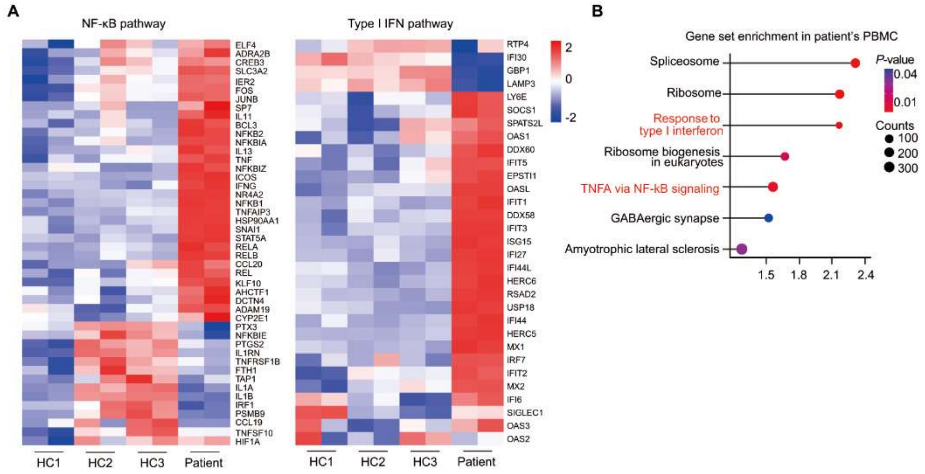


**Supplementary Figure 1. Transcriptional analysis of Inflammatory signaling in the patient**

(A) Expression level of genes on NF-κB (left) and type I IFN pathway (right) in PBMCs from the patient (P) and healthy controls (HC, n=3).

(B) Enrichment of upregulated differentially expressed genes in the patient's PBMC based on GSEA. NES, normalized enrichment score. *P* value is calculated by one-sided hypergeometric distribution test and adjusted by Bonferroni correction.

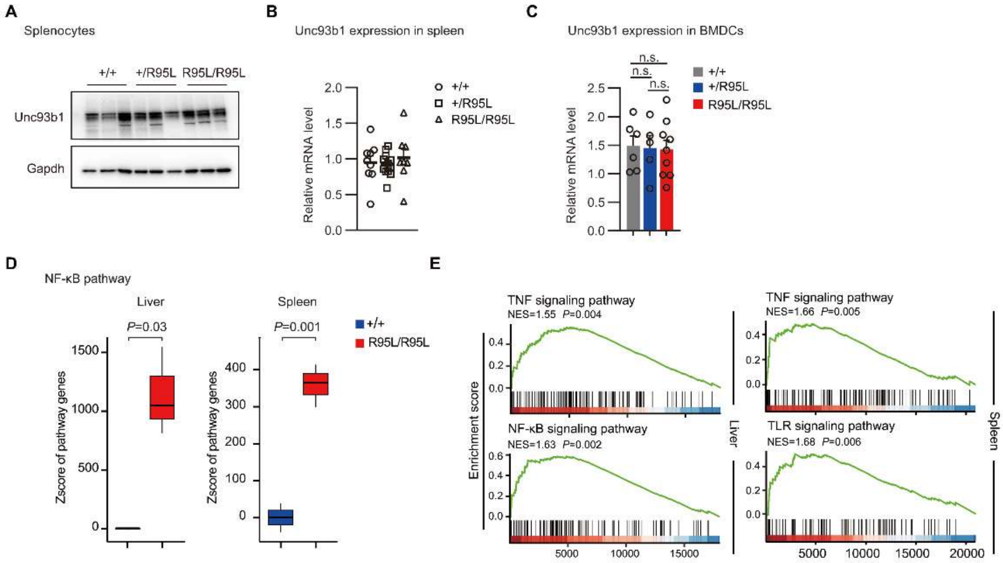


**Supplementary Figure 2. Transcriptional analysis of Inflammatory signaling in the liver and spleen from *Unc93b1*$^{R95L/R95L}$ mice**

(A) Immunoblotting of Unc93b1 in the splenocytes from *Unc93b1*$^{+/+}$, *Unc93b1*$^{+/R95L}$, and *Unc93b1*$^{R95L/R95L}$ mice.

(B) qPCR analysis of *Unc93b1* in splenocytes from *Unc93b1*$^{+/+}$ (n = 9), *Unc93b1*$^{+/R95L}$ (n = 12), and *Unc93b1*$^{R95L/R95L}$ (n = 8) mice.

(C) qPCR analysis of *Unc93b1* in BMDCs from *Unc93b1*$^{+/+}$ (n = 6), *Unc93b1*$^{+/R95L}$ (n = 5), and *Unc93b1*$^{R95L/R95L}$ (n = 9) mice. SEM and one-way ANOVA with Tukey's post hoc is used in statistics; n.s., non-significance.

(D) Comparison of Z-scores of indicated gene sets of NF-κB and type I IFN between the *Unc93b1*$^{R95L/R95L}$ (n = 3) and *Unc93b1*$^{+/+}$ mice (n = 3). Unpaired Student's *t* test is used in statistics. Boxes show the median (center line), 25th and 75th percentiles (upper and lower hinges), and whiskers extend to 1.5× the interquartile range.

(E) GSEA in indicated signaling pathway in liver (left) or spleen (right) tissues from the *Unc93b1*$^{R95L/R95L}$ and *Unc93b1*$^{+/+}$ mice. *P* value is calculated by one-sided hypergeometric distribution test and adjusted by Bonferroni correction. Permutation times of GSEA is set to 1000 as default.

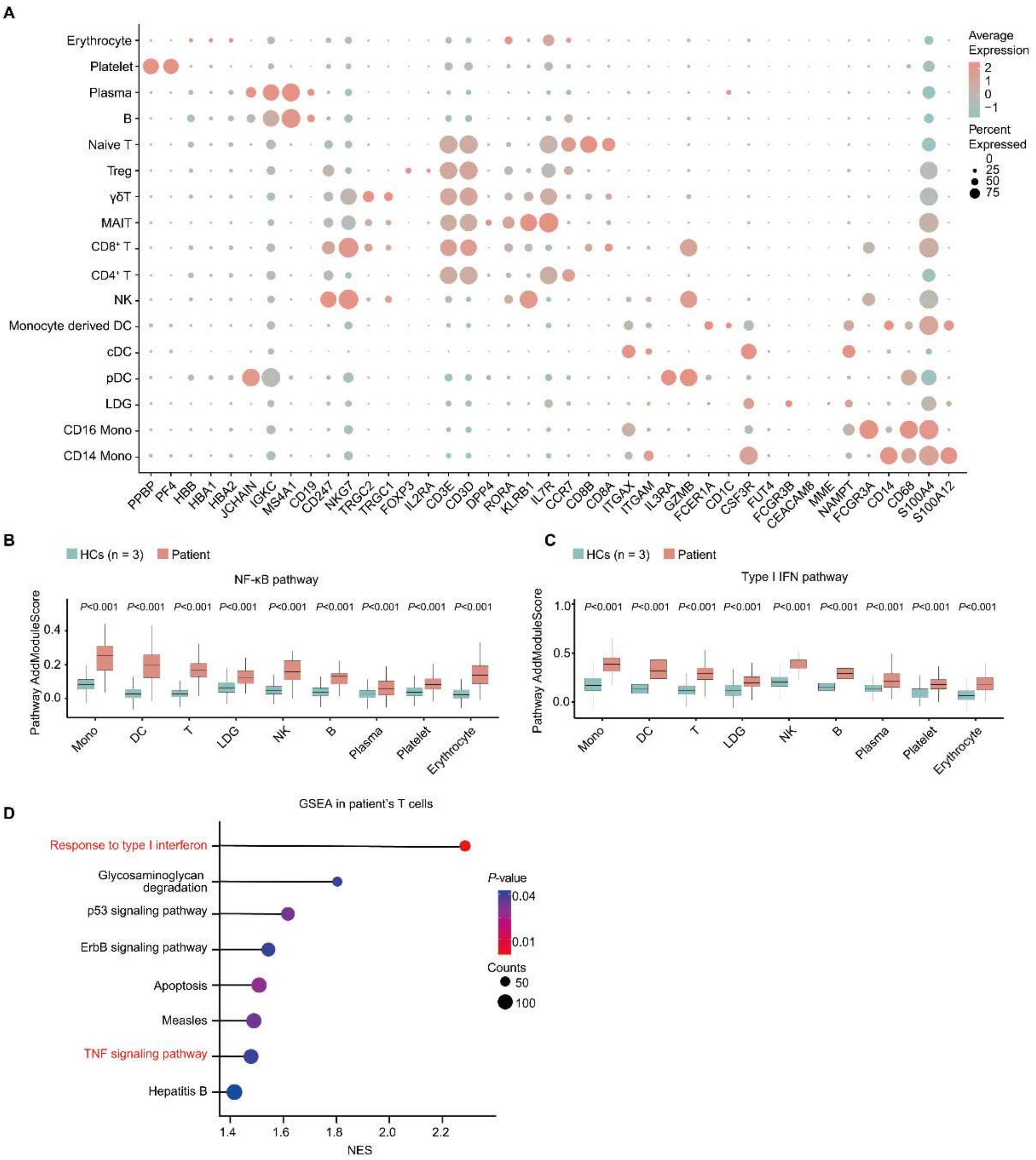


**Supplementary Figure 3. Single-cell transcriptional analysis revealed upregulated inflammatory signaling in the patient's PBMCs**

(A) Marker genes used for cluster annotation in single-cell RNA sequencing from the PBMCs of the patient and healthy controls.

(B,C) Boxplots of the score on NF-κB (B) and IFN response (C) pathways of cell subtypes based on Seurat AddModuleScore function corresponding to Fig. 3B. Unpaired Student's *t* test is used in statistics. Boxes show the median (center line), 25th and 75th percentiles (upper and lower hinges), and whiskers extend to 1.5× the interquartile range. Sample size for statistics calculation: Mono, ctrl: n = 3562, patient: n = 40; DC, ctrl: n = 302, patient: n = 412;

T, ctrl: n = 26222, patient: n = 1624; LDG, ctrl: 307, patient: n = 58; NK, ctrl: n = 1621, patient: n = 24; B, ctrl: n = 2950, patient: n = 22; Plasma, ctrl: n = 1610, patient: n = 104; Platelet, ctrl: n = 615, patient: n = 500.

(D) Enrichment of upregulated differentially expressed genes in T cells from the patient's PBMC based on GSEA. NES, normalized enrichment score. *P* value is calculated by one-sided hypergeometric distribution test and adjusted by Bonferroni correction.

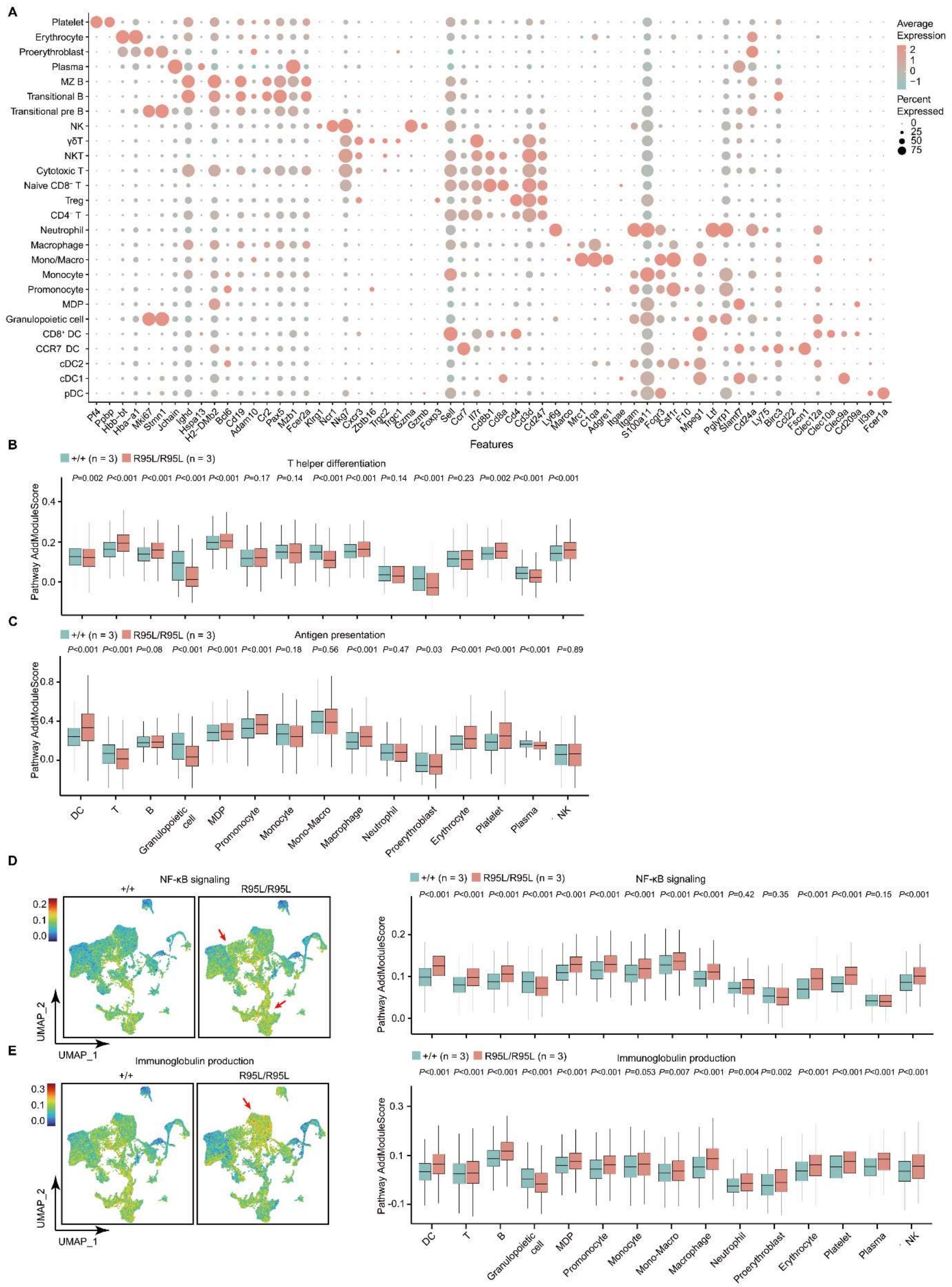


**Supplementary Figure 4. Single-cell transcriptional analysis in splenocytes from *Unc93b1*$^{R95L/R95L}$ and *Unc93b1*$^{+/+}$ mice**

(A) Marker genes used for cluster annotation in single-cell RNA sequencing of splenocytes from *Unc93b1*$^{R95L/R95L}$ and *Unc93b1*$^{+/+}$ mice.

(B,C) Boxplots of the score on T helper differentiation (B) and antigen presentation (C) pathways of cell subtypes based on Seurat AddModuleScore function. Unpaired Student's *t* test is used in statistics. Boxes show the median (center line), 25th and 75th percentiles (upper and lower hinges), and whiskers extend to 1.5× the interquartile range.

(D, E) Expression level on NF-κB pathway (D) and immunoglobulin production (E) pathways in *Unc93b1*$^{R95L/R95L}$ mice (n = 3), compared to *Unc93b1*$^{+/+}$ mice (n = 3). The score is calculated by the Seurat Addmodulescore method and mapped to the UMAP plot (left). Detailed statistical significance demonstrated with boxplots (right). Unpaired Student's *t* test is used in statistics. Boxes show the median (center line), 25th and 75th percentiles (upper and lower hinges), and whiskers extend to 1.5× the interquartile range.

Sample size for statistics calculation in panel B-E: DC, +/+: n = 3468, R95L/R95L: n = 4848; T, +/+: n = 19865, R95L/R95L: n = 11649; B, +/+: n = 14987, R95L/R95L: n = 12012; Granulopoietic cell, +/+: n = 275, R95L/R95L: n = 1248; MDP, +/+: n = 2303, R95L/R95L: n = 1732; Promonocyte, +/+: n = 1062, R95L/R95L: n = 3766; Monocyte, +/+: n = 805, R95L/R95L: n = 233; Mono-Macro, +/+: n = 1180, R95L/R95L: n = 616; Macrophage, +/+: n = 2017, R95L/R95L: n = 1374; Neutrophil, +/+: n = 207, R95L/R95L: n = 1074; γδT, +/+: n = 1482, R95L/R95L: n = 1032; NK, +/+: n = 2433, R95L/R95L: n = 888; Plasma, +/+: n = 1239, R95L/R95L: n = 1542; Proerythroblast, +/+: n = 207, R95L/R95L: n = 945; Erythrocyte, +/+: n = 463, R95L/R95L: n = 720; Platelet, +/+: n = 262, R95L/R95L: n = 464.

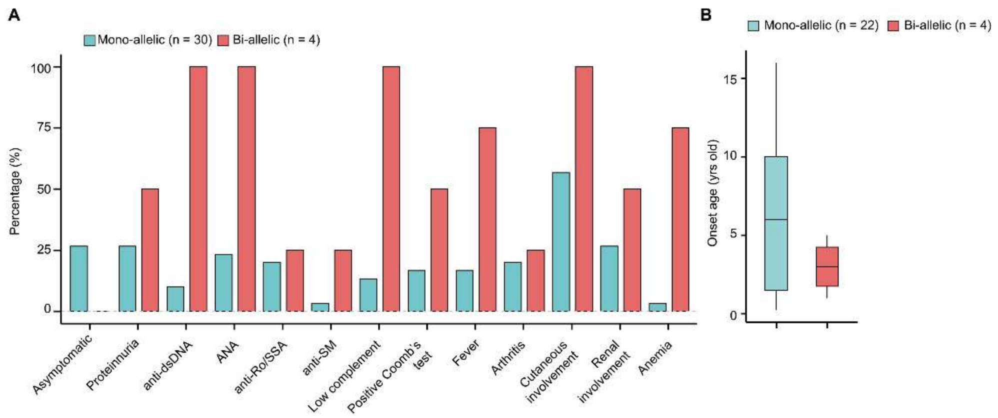


**Supplementary Figure 5.** The relation between the genotype and phenotype or onset age patients with of UNC93B1 GOF variants

(**A**) Observation of the percentage of indicated clinical manifestations in reported patients carrying mono-allelic (n = 30) or bi-allelic (n = 4) UNC93B1 GOF variant;

(**B**) The onset age of patients with mono-allelic (n = 22, excluded asymptomatic patient) or bi-allelic (n = 4) *UNC93B1* GOF variants. Boxes show the median (center line), 25th and 75th percentiles (upper and lower hinges), and whiskers extend to 1.5× the interquartile range
